# Mediation and Longitudinal Analysis to interpret the association between clozapine pharmacokinetics, pharmacogenomics, and absolute neutrophil count

**DOI:** 10.1101/2023.08.22.23294262

**Authors:** Siobhan K. Lock, Sophie E. Legge, Djenifer B. Kappel, Isabella R. Willcocks, Marinka Helthuis, John Jansen, James T. R. Walters, Michael J. Owen, Michael C. O’Donovan, Antonio F. Pardiñas

**Author notes:** Corresponding Author: Antonio F. Pardiñas, Centre for Neuropsychiatric Genetics and Genomics, Division of Psychological Medicine and Clinical Neurosciences, Hadyn Ellis Building, Maindy Road, Cardiff University, Cardiff, UK, CF24 4HQ.

## Abstract

Clozapine is effective at reducing symptoms of treatment-resistant schizophrenia, but it can also induce several adverse outcomes including neutropenia and agranulocytosis. We used linear mixed-effect models and structural equation modelling to determine whether pharmacokinetic and genetic variables influence absolute neutrophil count in a longitudinal UK-based sample of clozapine users not currently experiencing neutropenia (N = 811). Increased daily clozapine dose was associated with elevated neutrophil count, amounting to a 133 cells/mm^3^ rise per standard deviation increase in clozapine dose. One-third of the total effect of clozapine dose was mediated by plasma clozapine and norclozapine levels, which themselves demonstrated opposing, independent associations with absolute neutrophil count. Finally, CYP1A2 pharmacogenomic activity score was associated with absolute neutrophil count, supporting lower neutrophil levels in CYP1A2 poor metabolisers during clozapine use. This information may facilitate identifying at-risk patients and then introducing preventative interventions or individualised pharmacovigilance procedures to help mitigate these adverse haematological reactions.

## Introduction

Antipsychotics are the primary pharmacological treatments for people with schizophrenia. Response to these drugs is highly variable, and approximately one-third of patients respond insufficiently after several prescriptions ^1^. Formally, the term “treatment-resistant schizophrenia” applies to individuals who do not respond to at least two different antipsychotics taken at a therapeutic dose, for an appropriate length of time, and after having ruled out non-compliance ^2,3^. Clozapine is the sole evidence-based pharmacotherapy for treatment-resistant schizophrenia, but some clinicians are hesitant to prescribe it due to a range of potential adverse drug reactions (ADRs). The best known of these involve a decline in absolute neutrophil count (ANC) that leads to neutropenia and ultimately agranulocytosis ^4^. Agranulocytosis is a rare, severe, and potentially lethal clozapine-induced ADR that is currently unpredictable ^5^. However, a less severe decline in ANC may also be problematic; it has been suggested that this can result in partial suppression of the immune system even before formal criteria for neutropenia are met, increasing vulnerability to infectious diseases ^6^.

The mechanisms underlying clozapine-induced neutrophil loss are unknown, but it is thought to arise through processes involving clozapine metabolites. The CYP family of enzymes, notably CYP1A2, CYP2D6, and CYP3A4, are heavily involved with the biotransformation of clozapine through its metabolic pathway^7^, leading to norclozapine and clozapine-N-oxide as key products. However, the drug can also be oxidised to a nitrenium ion, a nitrogenous intermediate characterised by its high reactivity and ability to bind with cells^8^. The conversion of clozapine to the nitrenium ion is mediated by neutrophil action. Activated neutrophils combat infection by producing an antimicrobial agent, hypochlorous acid, via the enzyme myeloperoxidase. Both hypochlorous acid and myeloperoxidase may also react with clozapine to form the intermediate nitrenium ion^9,10^. It is thought that this reactive intermediate may harm neutrophils through two primary mechanisms: haptenation, in which the nitrenium ion binds irreversibly to neutrophil cell surface proteins, or through overactivation of the glutathione system which may be recruited to form conjugates with the nitrenium ion and detoxify it. Indeed, it is known that both these mechanisms can lead to neutrophil apoptosis ^11–13^.

Regular blood monitoring is a requirement of clozapine prescription both in the UK and in many other countries to reduce the risks of progression from low neutrophil count to formal agranulocytosis ^14^. Nevertheless, despite the superiority of clozapine for managing treatment-resistant schizophrenia in comparison to standard first-line antipsychotics, concerns about ADR risk and monitoring requirements are primary contributors to this drug being underutilised worldwide ^15^. For this reason, identifying factors that are predictive of low neutrophil counts in an otherwise healthy sample of clozapine users could help clinicians to improve clozapine use while supporting patient safety and wellbeing. For example, if clozapine users susceptible to increased risk for infections could be identified, potential harm might be mitigated by prioritising them for seasonal vaccinations or making changes to their blood monitoring regime.

Past research ^16–18^ explored associations between ANC and daily clozapine dose as well as plasma concentrations of clozapine and norclozapine. Generally, higher plasma clozapine concentration has been found to be associated with lower ANC, whereas higher plasma norclozapine concentration has been reported to be associated with higher neutrophil counts. However, as highlighted in a recent review ^19^, there is inconsistency in the literature regarding both the direction and magnitude of effects observed. While this could be in part due to differences in statistical methodology and the use of small samples, it could equally reflect the idiosyncrasy of the ADR leading to ANC decline, alongside the challenges of disentangling the impact of clozapine and its related variables from genetic, demographic, or lifestyle factors that may also influence neutrophil counts.

The present research aims to replicate and extend these previous studies by exploring predictors of ANC in a large, UK-based sample of clozapine users with TRS in whom longitudinal measures were available for both pharmacokinetic variables and full blood counts. Linear mixed-effect models (LMMs) were used to explore associations between pharmacokinetic and pharmacogenomic variables and ANC, while accounting for between- and within-individual variability. Following this, a Structural Equation Modelling (SEM) framework was used to further disentangle the contributions of clozapine dose, from plasma clozapine and plasma norclozapine levels. Genetic predictors relating to clozapine and norclozapine metabolism, as well as baseline variation in ANC were also investigated to determine whether they influenced neutrophil counts in our sample.

## Methods

### Sample collection / Participants

We used Full Blood Counts (FBC) and pharmacokinetic assay data from the CLOZUK3 sample ^20^, the most recent wave of the CLOZUK study ^21^. All participants had a diagnosis of treatment-resistant schizophrenia and were prescribed clozapine. Samples were anonymously collected in the UK from the Zaponex Treatment Access System (ZTAS), a clozapine monitoring framework managed by Leyden Delta B.V. (Nijmegen, Netherlands). Sample collection and data extraction procedures for CLOZUK have been detailed previously ^21,22^. The CLOZUK study received UK National Research Ethics Service approval (reference 10/WSE02/15), in accordance with the requirements of the UK Human Tissue Act 2004.

### Inclusion/Exclusion criteria

CLOZUK3 contains longitudinal assay data from participants older than 18 years of age who had not been previously included in prior waves of CLOZUK. We excluded data from assays with missing FBC or pharmacokinetic information or with clerical errors (e.g., several assays taken at the same date and time yielding different results) from further analyses. To retain only apparently healthy individuals, the FBC data were curated by removing any assay showing an ANC outside of the normal reference range (2000 – 7500 cells/mm^3^). Additionally, and as in previous research ^20^, we removed pharmacokinetic assays (i) where the gap between clozapine intake and blood sampling was outside a 6-24 hour window; (ii) where clozapine and norclozapine plasma concentrations were below instrument detection levels (< 0.05 mg/L); (iii) showing a low daily clozapine dose (< 100mg, potentially indicative of drug titration) or (iv) where the metabolic ratio suggested non-adherence (< 0.5 or > 3.0).

### Genetic Data

A subset of individuals (N = 523) in the sample had linked genetic data, based on genotypes from an Illumina Infinium Global Sequencing Array-24 (Illumina Inc, USA). Details regarding the curation and imputation of this genetic data have been described elsewhere ^20^ and summarised in the **Supplementary Note**. Pharmacogenomic star alleles (i.e., genetic variants or combinations of variants constituting pharmacogenomic markers) for CYP1A2 were called using PyPGx v0.20.0 ^23^ on the imputed array data. Enzyme activity scores inferred from these star alleles were included in LMMs to determine whether genetic predictors of CYP1A2 activity were associated with ANC. Other pharmacogenomic SNPs (Table 1) identified in a GWAS of clozapine metabolism ^24^ and included in a previous analysis exploring predictors of ANC ^18^ were also investigated.

**Table 1.** SNPs included in regression analyses exploring the impact of pharmacogenomic variation neutrophil levels. SNP = Single Nucleotide Polymorphism; CHR = Chromosome; ALT = Alternative (or Minor) allele.

| SNP | CHR | Gene | ALT | Main Association |
| --- | --- | --- | --- | --- |
| rs2011425 | 2 | UGT1A4 | G | Alternative allele linked with decreased plasma norclozapine levels <sup>24</sup>. |
| rs61750900 | 4 | UGT2B10 | T | Alternative allele linked with decreased plasma norclozapine levels <sup>24</sup>. |
| rs1126545 | 10 | CYP2C18 | T | Alternative allele linked with increased metabolic ratio <sup>24</sup>. |
| rs2472297 | 15 | CYP1A1/CYP1A2 | T | Alternative allele linked with decreased plasma clozapine levels <sup>24</sup>. |
| rs2814778 | 1 | ACKR1 | C | Alternative allele homozygosity (Duffy-Null genotype) linked with non-pathological baseline ANC <sup>26</sup>. |

Polygenic Scores (PGS) for clozapine and norclozapine metabolism were calculated via PRSice2 v2.35 ^25^ as part of a previous study ^22^. These were included in secondary analyses. We also explored the impact of the Duffy-null genotype (rs2814778; C/C homozygote) as this has been associated with decreased ANC in clozapine users of African, Asian, and Middle Eastern ancestries ^26^. Finally, the Human Leukocyte Antigen (HLA) system has long been understood as a crucial component of the immune system, with past work linking this genetic locus to agranulocytosis in clozapine users ^27^. Therefore, we imputed HLA types using HIBAG v1.34.1 ^28^ and incorporated these into LMMs to explore their impact on ANC. Detailed descriptions of CYP1A2 pharmacogenomic allele calling and HLA genotype imputation are found in the **Supplementary note**.

### Statistical Analysis

A Directed Acyclic Graph (DAG) was first drawn to consider the possible causal structure of the clozapine dose – ANC relationship ^29^. As previously recommended ^30^, we explicitly report the rationale for included DAG nodes and relationships in Supplementary Table 3.

Data analyses were performed in R v4.1.1 using R Studio 2023.06.1+524 ^31^. The longitudinal dataset was analysed using LMMs in *lme4* with ANC as the outcome variable. A baseline model, in line with previous work^18^, included three pharmacokinetic variables related to clozapine and its metabolism (i.e., daily clozapine dose, plasma clozapine concentration, and plasma norclozapine concentration) alongside covariates for age, age^2^, sex, and the time between the dose intake and blood sampling (TDS). Participant ID was included in these models as a random effect term. All predictor variables were standardised as described in the **Supplementary Note**, before fitting the regression model. Further analyses included pharmacogenomic variables (i.e., CYP1A2 activity scores, and the genotypes of pharmacogenomic SNPs outlined in Table 1).

Secondary analyses extended the LMMs by testing for associations between the additional genetic predictors (i.e., PGS for clozapine and norclozapine metabolism, the Duffy-null genotype, and HLA genotypes) with neutrophil counts.

Due to the difficulty of implementing and interpreting causal analyses on longitudinal datasets with irregular time points ^32^, mediation models were fit using the lowest value of ANC reported for each individual as the outcome variable. SEM was conducted using *lavaan* ^33^ including clozapine and norclozapine plasma concentrations as mediating variables, daily clozapine dose as the exposure, and lowest ANC as the outcome. Residualised versions of these variables were included in the model, as described in the **Supplementary Note**. Predictor variable residuals were standardised before inclusion in the model.

Further sensitivity analyses tested the robustness of these mediation models, assessing both the impact of using residualised variables and using cross-sectional, as opposed to longitudinal data. Single-mediator analyses were also implemented in the *mediation* R package ^34^. While this approach can estimate direct and indirect effects in longitudinal datasets it can only accommodate a single mediator variable. Therefore, it was not appropriate for the primary analysis of the multiple mediation model that we defined and evaluated using SEM on cross-sectional data in *lavaan*. Finally, we attempted to formally replicate the analyses described in previous studies ^17,18^ by implementing linear models, as reported in the **Supplementary Note**.

## Results

The final curated CLOZUK3 longitudinal dataset included 811 participants, with a total of 2,362 FBC and pharmacokinetic assays taken on the same day. Participants had a mean (SD) age of 40.1 (12.2) years; 28.9% were female (N = 234) and 71.1% were male (N = 577). Full descriptive statistics of the sample at the point of lowest ANC are given in Table 2. While the first occurrence of each individual on our ZTAS dataset is not necessarily the date they started clozapine, we note that about a third of the CLOZUK3 individuals with valid data (32.6%; 264/811) had records spanning at least a year of clozapine treatment.

**Table 2.**
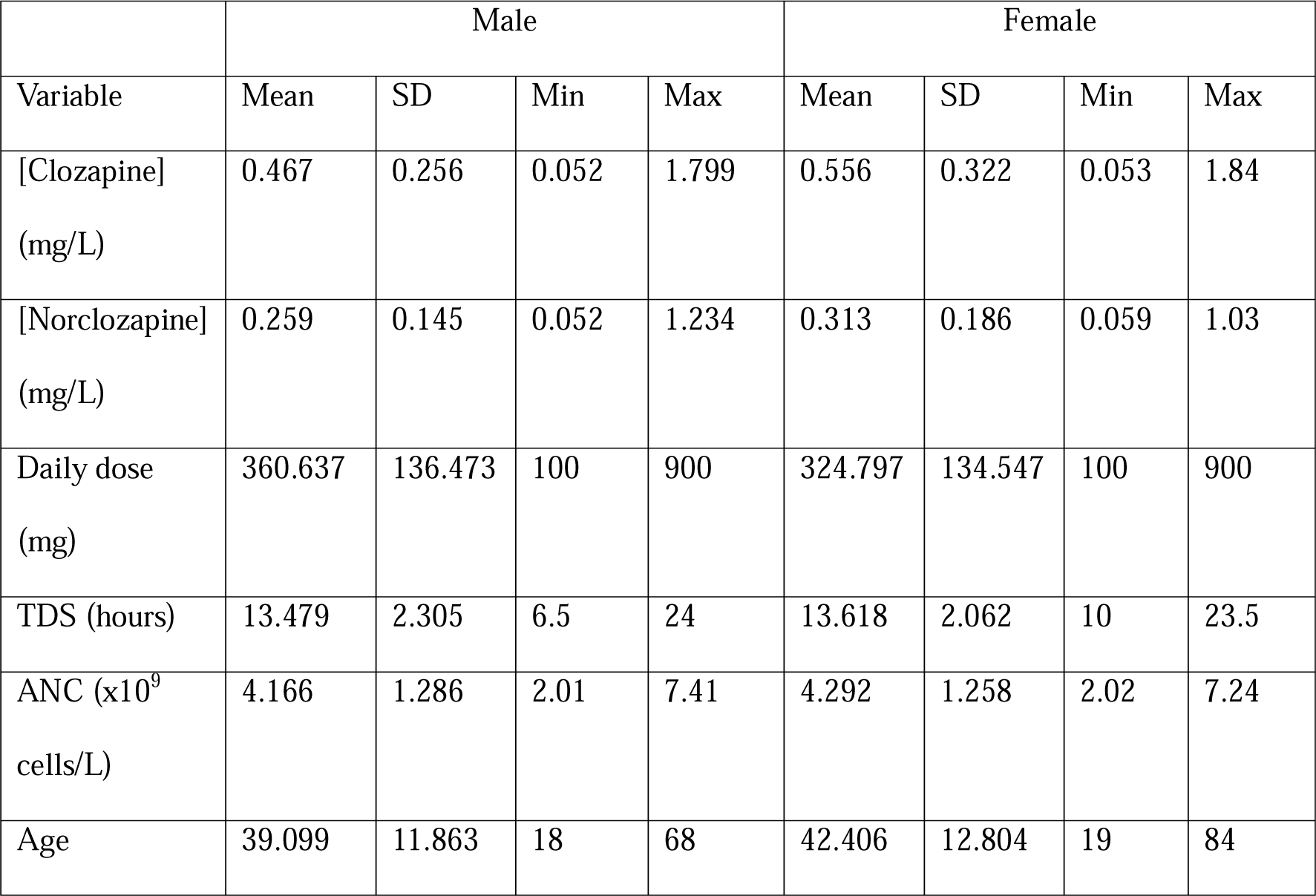
Summary of CLOZUK3 variables used in mediation analysis. Descriptive statistics presented for a ‘cross-sectional’ version of CLOZUK3 where only the entry associated with the lowest value of ANC per person is retained. TDS = Time between Dose and Sample; ANC = Absolute Neutrophil Count; [Clozapine] = Clozapine plasma concentration; [Norclozapine] = Norclozapine plasma concentration.

|  | Male |  |  |  | Female |  |  |  |
| --- | --- | --- | --- | --- | --- | --- | --- | --- |
| Variable | Mean | SD | Min | Max | Mean | SD | Min | Max |
| [Clozapine]<br>(mg/L) | 0.467 | 0.256 | 0.052 | 1.799 | 0.556 | 0.322 | 0.053 | 1.84 |
| [Norclozapine]<br>(mg/L) | 0.259 | 0.145 | 0.052 | 1.234 | 0.313 | 0.186 | 0.059 | 1.03 |
| Daily dose<br>(mg) | 360.637 | 136.473 | 100 | 900 | 324.797 | 134.547 | 100 | 900 |
| TDS (hours) | 13.479 | 2.305 | 6.5 | 24 | 13.618 | 2.062 | 10 | 23.5 |
| ANC ($\times 10^9$<br>cells/L) | 4.166 | 1.286 | 2.01 | 7.41 | 4.292 | 1.258 | 2.02 | 7.24 |
| Age | 39.099 | 11.863 | 18 | 68 | 42.406 | 12.804 | 19 | 84 |

The DAG (Supplementary Figure 2) displays the possible causal paths between ANC (the outcome), daily clozapine dose (the exposure variable), and plasma concentrations of clozapine and norclozapine (potential mediators between dose and ANC).

### Significant associations between pharmacokinetic and pharmacogenomic variables with ANC

All pharmacokinetic variables were significantly associated with ANC (Table 3). ANC was inversely associated with clozapine plasma concentration (β = −0.166; *p* = 0.002) and positively associated with norclozapine plasma concentration (β = 0.219; *p* = 6.06 × 10^-5^). In the original FBC scales, a reduction in ANC of 166 cells/mm^3^ was observed for every standard deviation increase in plasma clozapine concentration. Likewise, each standard deviation increase in plasma norclozapine concentration was accompanied by a 219 cells/mm^3^ increase in ANC. We note that daily clozapine dose was also associated with ANC in this model (β = 0.133, *p* = 1.08 × 10^-4^), corresponding to an estimated increase of 133 cells/mm^3^ per standard deviation increase in the daily dose.

**Table 3.**
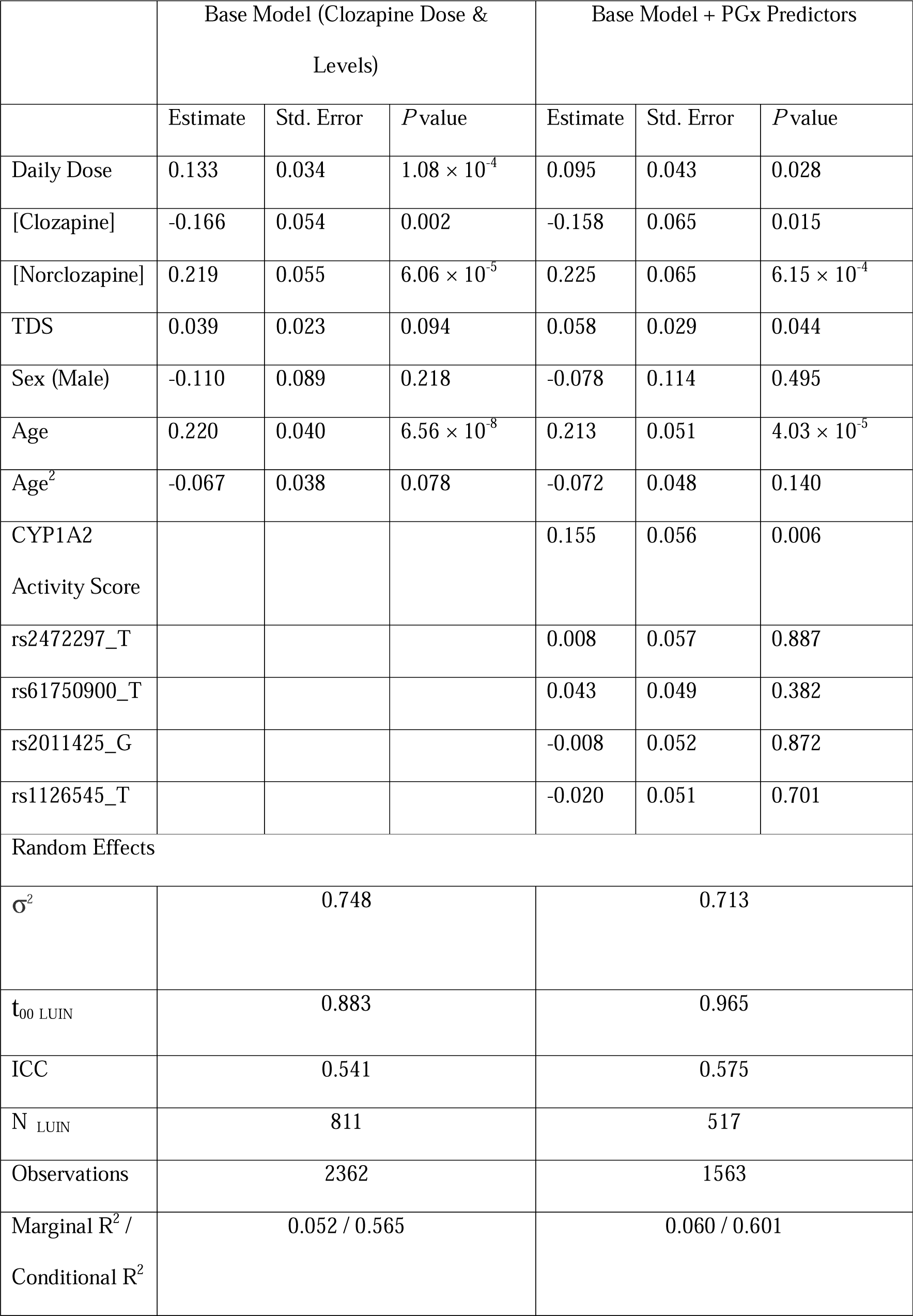
Results of two Linear Mixed-Effect Models exploring predictors of Absolute Neutrophil Count (ANC). Standardised regression coefficients are reported alongside standard error and *p* values estimated using the *lmerTest* package. PGx = Pharmacogenomic; TDS = Time between Dose and Sample; [Clozapine] = Clozapine plasma concentration; [Norclozapine] = Norclozapine plasma concentration; LUIN = Participant Identifier used in CLOZUK3; σ^2^ = Residual Variance; ICC = Intraclass Correlation Coefficient; t_00 LUIN_ = Random Intercept Variance; N _LUIN_ = Number of participants.

The pharmacokinetic variables dose, clozapine plasma concentration, and norclozapine plasma concentration remained significantly associated with ANC after incorporating pharmacogenomic predictors in the model for the subset of individuals with genetic data (Table 3). We found no evidence for association between ANC and any of the pharmacogenomic SNPs, PGS for clozapine and norclozapine, or variation in the HLA region **(Supplementary Note)**.

We saw a significant, positive association between CYP1A2 activity score and ANC, in which increased CYP1A2 function (i.e., rapid CYP1A2 metabolism), was associated with increased neutrophil counts (β = 0.155; *p* = 0.006). As described in the **Supplementary note**, this association was independent of rs2472297, a known regulator of CYP1A2 activity and a genome-wide significant SNP in clozapine pharmacokinetic GWAS ^24^.

Finally, the presence of the Duffy-Null genotype, observed in just under 5% of the CLOZUK3 sample, was significantly associated with reduced ANC (β = −0.770; *p* = 0.002).

### Plasma Clozapine and Norclozapine levels mediate the Dose – ANC association

The primary model (Figure 1) showed evidence of a significant direct effect of daily clozapine dose on ANC (β = 0.150, *p* = 8.87 x 10^-4^). The indirect path via both clozapine and norclozapine plasma concentration was also significant (β = 0.057, *p* = 0.018). However, no indirect effect was observed when plasma clozapine concentration was considered as the sole mediator (β = −0.028, *p* = 0.116). Secondary analyses revealed that CYP1A2 activity scores appeared to account for part of these associations (**Supplementary note**). However, as only some of the CLOZUK3 sample was genotyped (523/811), these models would have reduced statistical power in relation to our main analyses and their results should be considered with caution.

**Figure 1.**
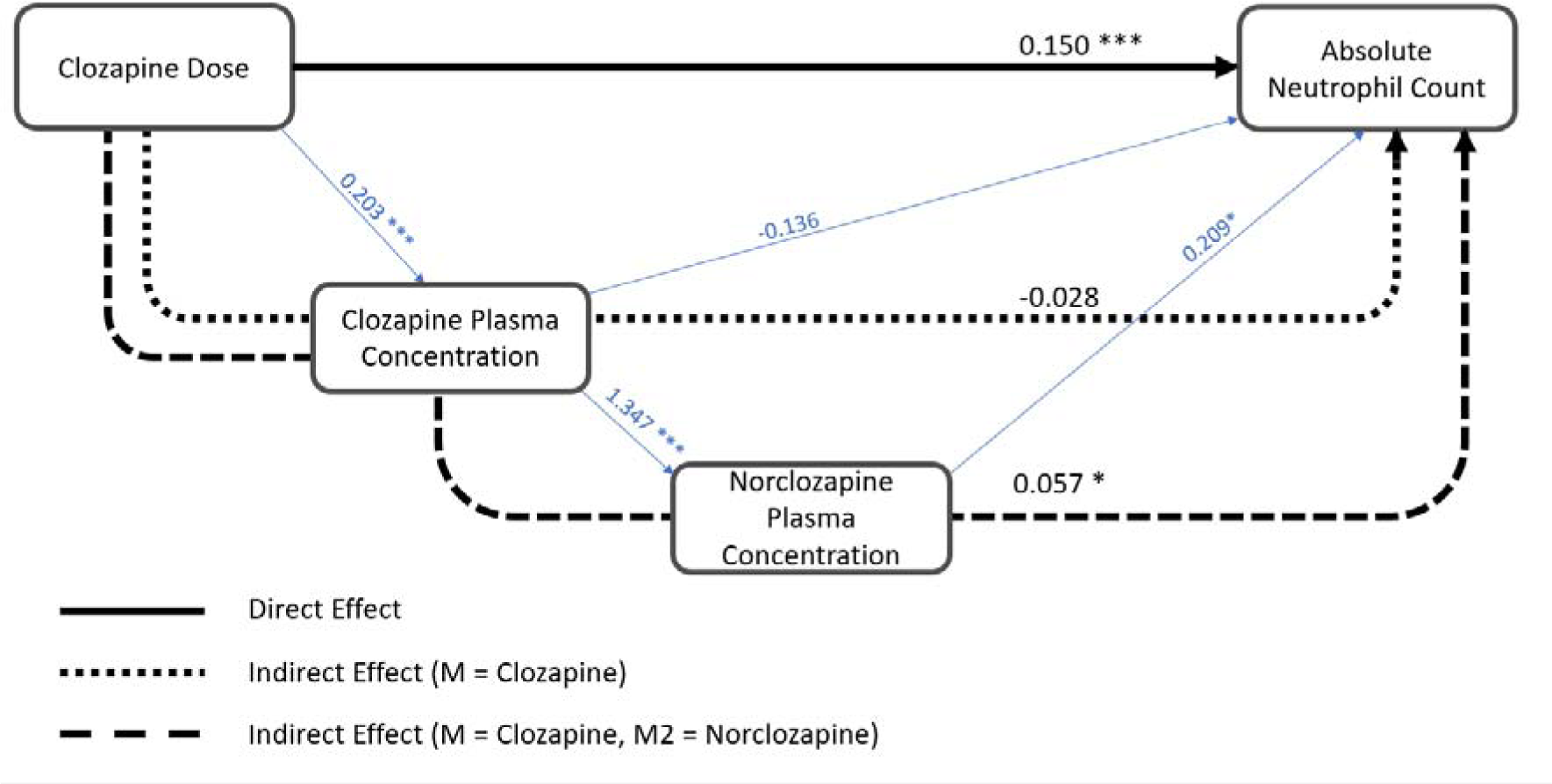
Path diagram showing association between Clozapine Dose and Lowest Absolute Neutrophil Count with Plasma Clozapine concentration and Plasma Norclozapine concentration as mediators. Plot edges are labelled with standardised regression coefficients. Variables included in Structural Equation Model are residualised versions of parent measures. Associations between model variables are shown in blue, whereas the overall direct and indirect paths are shown in black. M = Mediator. * p<0.05 ** p<0.01 *** p<0.001

Sensitivity analyses were performed using non-residualised variables in the model, and also by testing single mediators in the longitudinal dataset. These were all consistent with the results of the primary analyses, suggesting that our models were not compromised through the covariates considered for residualisation, or the cross-sectional nature of our multiple mediation tests (**Supplementary note**).

## Discussion

### Key Findings

The results of the present study provide evidence for associations between both pharmacokinetic and genetic variables with neutrophil count in the CLOZUK3 sample. Daily clozapine dose was positively associated with ANC, and approximately a third of its overall impact was mediated by plasma clozapine and norclozapine levels. We also observed opposing effects of plasma clozapine and norclozapine concentrations on ANC, with plasma clozapine levels inversely associated, and plasma norclozapine levels positively associated with neutrophil counts. Finally, we found evidence that both CYP1A2 activity score and the Duffy-null genotype were associated with ANC.

The direct, positive association between clozapine dose and ANC across our analyses is both novel and unexpected, given that past research has found clozapine dose to be a poor predictor of ANC ^18,35^. Furthermore, the direction of the effect is inconsistent with expectations, given that clozapine is believed to induce neutropenia or agranulocytosis ^36^. This positive relationship could reflect an immune response to clozapine resulting in elevated neutrophil counts, as reported in rats ^37^ and humans ^38^. Further work has shown that an increase in immature neutrophils may occur as part of this immune response ^39^, which could result in raised ANC. Alternatively, the positive association may represent reverse causation, through clinicians altering clozapine prescriptions in response to the full blood count results in ways that are not explicitly endorsed by treatment guidelines. For example, some clinicians might aim to counteract a patient’s low neutrophil levels by reducing daily clozapine dose in hopes of avoiding discontinuation. Alternatively, they might become reluctant to increase clozapine dose due to fear of prompting further neutrophil loss.

The associations between plasma clozapine and norclozapine levels with ANC are consistent with past research ^16–18^. This work cannot firmly establish which aspect of clozapine use engenders neutrophil loss as we have not tested the full range of secondary and tertiary metabolites of this drug (such as N-oxide or N-glucuronides), some of which are known to have reactive properties ^40^. However, taken together these results suggest that individuals with high clozapine levels may be prone to displaying low ANC, and that the plasma norclozapine concentration is unlikely to reflect the toxic component of this process.

The present study also explored the impact of a well-established pharmacogenomic variable, CYP1A2 enzyme activity as inferred from classic star allele calling. We observed a positive association between the CYP1A2 activity score and ANC, in which increased enzyme activity is associated with greater neutrophil counts or, conversely, poor metabolism is associated with lower ANC. This is consistent with current knowledge about clozapine’s metabolic pathway. Both CYP1A2 and CYP3A4 are involved in the metabolism of clozapine, either producing norclozapine or to a lesser extent, clozapine-N-oxide ^24^. Therefore, increased CYP1A2 activity should result in the faster conversion of clozapine to norclozapine, reducing the potentially toxic impact of other metabolites (including clozapine itself) on neutrophil counts. Thus, we provide preliminary evidence that genetically-inferred enzyme activity may have utility for predicting neutrophil counts in a non-neutropenic, clozapine-using population. Future research should be performed across different samples to ascertain the robustness of these results, and following this, could be extended to explore other enzymes involved in clozapine metabolism. It would also be of interest to determine whether similar associations occur in patients with low neutrophil counts indicative of neutropenia and agranulocytosis.

In contrast to previous work ^18^, none of the included pharmacogenomic SNPs were associated with ANC. While we caution this could be partly due to the limited size of the sample with genetic data (N = 523), it does reinforce the need for more genomic studies to better understand the possible impact of these variants on neutrophil counts in clozapine users. Furthermore, neither the PGS for clozapine and norclozapine metabolism nor the assessed HLA genotypes were significantly associated with ANC. However, before correction for multiple comparisons several HLA alleles (i.e., *DRB1*16:01, DRB1*04:04, DRB1*01:03*) were nominally significant. While increased frequency of *HLA-DRB1*16:01* has been previously associated with clozapine-induced agranulocytosis cases ^41^, it has not yet been linked to neutrophil levels in a non-neutropenic population. The other nominally associated alleles have not been implicated in clozapine-induced neutrophil loss but do provide direction for further work investigating the impact of this locus on neutrophil levels in clozapine users.

Finally, as expected from previous work^26^, a genetic predictor associated with lower non-pathological baseline neutrophil counts, the Duffy-Null genotype, was negatively associated with ANC. This is consistent with observations that the genotype is associated with reduced non-pathological neutrophil counts^26^. This genotype is most common in individuals of African ancestries but may also be present in those of Middle Eastern and Asian ancestries as it confers resistance to malaria ^42^. However, our work confirms that the influence of this genotype on ANC is also apparent in a sample of people with primarily European ancestries. This supports the notion that testing patients for this genotype might be more helpful in interpreting their blood monitoring assays than simply considering ethnicity as a driver of differences in ANC ^43^. Such genetic testing could increase clinician confidence when prescribing clozapine to people of ancestries where this genotype is common and thus help to combat disparities in clozapine use and prescription and widen access to this medication ^44^.

### Strengths and Limitations

In the context of research on the pharmacogenomics of treatment-resistant schizophrenia, CLOZUK3 is a relatively large sample linked to an extensive longitudinal blood monitoring dataset. It is therefore better powered than most previous studies to assess the relationships between clozapine dose and metabolites on ANC. Data availability also allowed us to merge and analyse FBC and pharmacokinetic assays taken on the same day, providing a precise relationship between neutrophil counts and plasma clozapine and norclozapine levels. To our knowledge, this is also the first work to establish associations between pharmacogenomics-inferred CYP1A2 activity with neutrophil counts in clozapine users.

A limitation of the present work is that the CLOZUK3 sample was not fully genotyped. Nevertheless, the key pharmacokinetic associations observed in the LMM remain when explored in the subset of the genomically informative sample (**Supplementary Note**). Furthermore, the participants in the CLOZUK3 dataset were primarily of European ancestry limiting the extent to which the findings can be generalised to other populations. Our findings therefore need to be tested in non-European individuals, as exemplified by the strength of known ancestry-specific genetic effects on ANC^26^.

We were unable to account for the effects of concomitant medication (e.g., oral contraceptives, some antidepressants) or lifestyle factors (e.g., caffeine consumption, cigarette smoking) that can modify enzyme activity and thus impact clozapine metabolism ^45–47^. Moreover, there is some evidence that cigarette and caffeine consumption can be associated with white blood cell counts ^48–50^, and information about these variables was similarly unavailable. Therefore, our study needs replication in an independent sample where the potential of substance use to either influence clozapine metabolism via phenoconversion or act as a confounder in our models can be explicitly tested.

### Implications

Clozapine use is associated with decreased all-cause mortality compared to other commonly used antipsychotic drugs ^51^; however unexpected deaths by various causes remain a rare and currently unpredictable feature of the medication ^52^. While ANC is not necessarily related to mortality itself, a progressive depletion of immune cells has been argued to be a primary contributor to the susceptibility to infectious disease exhibited by clozapine users ^53,54^. The present work found several associations between pharmacokinetic and genetic variables with ANC in a UK-based sample of clozapine users with no detectable immune-related ADRs. Our results could have clinical applicability from the perspective of treatment management, supporting the view that clozapine dose might become a modifiable risk factor in cases with abnormal neutrophil counts. While agranulocytosis and severe neutropenia are considered dose-independent, subclinical variation in ANC might respond to dose alterations. Clozapine metabolism is also a factor to consider as plasma clozapine and norclozapine levels were also significantly associated with ANC in our sample. However, large inter- and intra-individual differences in levels at fixed doses might make it complicated to influence these variables in practice ^55^, though they still could find applicability for the identification of patients at risk of extreme ANCs.

This work adds to a body of research aiming towards a complete understanding of the factors that influence ANC in clozapine users, which could have value in improving access to this gold-standard medication. Currently, there is reluctance amongst some clinicians to prescribe clozapine to patients with schizophrenia. This is primarily due to the risk of neutropenia and agranulocytosis, and the accompanying need for therapeutic blood monitoring to ensure patient safety. While effective at reducing deaths from clozapine-induced agranulocytosis in the UK ^56^, haematological monitoring is time-consuming both from the perspective of the patient, and the clinician. Appropriately interpreting the effects of pharmacokinetic and genetic variables that influence neutrophil loss might allow the design of stratification strategies for clozapine users based on their likelihood of immune-related ADRs, with appropriate adjustments of prescription and monitoring. This could also inform preventative interventions targeting those most likely to experience neutrophil loss, and subsequent immune decline, which may help to prevent serious illness over the duration of clozapine use. Some examples of this might include encouraging at-risk patients to take up offers of seasonal vaccinations, particularly given evidence of reduced vaccine uptake in people with psychiatric disorders ^57^; testing markers of inflammation and adjusting treatment, or introducing further mitigations accordingly. Knowledge of these variables could act as an additional layer of information to guide clinical decision-making and ultimately help to widen access to clozapine via a two-pronged approach: enhancing safety for those at highest risk while reducing obstacles to treatment use for those at lowest risk.

### Conclusions

Here, we show daily clozapine dose was positively associated with ANC, with clozapine pharmacokinetics (indexed by clozapine and norclozapine plasma levels) accounting for a third of the total effect. Our analysis of multiple pharmacokinetic and pharmacogenomic variables supports and expands on the results from past research, which for decades has suggested an opposing relationship between ANC and plasma clozapine and norclozapine levels. We build on this to show that these effects exist in a sample in which about a third of the individuals were taking clozapine for over a year, commonly considered to be at a lower risk of immune-related ADRs.

The pharmacokinetic analysis was supplemented by genetic covariates, notably a CYP1A2 activity score inferred from pharmacogenomic star alleles. CYP1A2 activity was positively associated with ANC; however, no associations were seen between neutrophil counts with pharmacogenomic SNPs previously associated with clozapine metabolism. Additional work in larger, more genetically diverse samples is required to clarify the role of pharmacogenomic variation in clozapine metabolism, and its capacity to influence neutrophil levels in clozapine users. In all, this work advances our understanding of the impacts of clozapine use on neutrophil counts, which in the future may help to improve access to clozapine via the development of targeted interventions and personalised drug monitoring schedules based on individual risk factors.

## Data Availability

Code for reproducing all the main analyses in R is available online https://locksk.github.io/clozuk3-anc/. Additional scripts for calling pharmacogenomic and HLA alleles are available at https://github.com/locksk/clozapine-predictors-of-anc/. To comply with the ethical and regulatory framework of the CLOZUK project, access to individual-level data requires a collaboration agreement with Cardiff University. Requests to access deidentified datasets, data dictionaries, and other summaries from the CLOZUK project should be directed to James Walters.

https://github.com/locksk/clozapine-predictors-of-anc/

## Acknowledgements

We acknowledge the support of the Supercomputing Wales project, partly funded by the European Regional Development Fund (ERDF) via the Welsh Government. We thank Joanne Morgan, Lesley Bates, Catherine Bresner and Lucinda Hopkins (Cardiff University) for laboratory sample management. We also thank Adrian King (Magna Laboratories) for contributing to the sample collection and data preparation of CLOZUK3.

