## Supplementary Notes for "Mediation and Longitudinal Analysis to interpret the association between clozapine pharmacokinetics, pharmacogenomics, and absolute neutrophil count"

**Supplementary Information**

### Supplementary Notes: Genetic Variables

#### Quality Control of Genetic data

A subset of the CLOZUK3 sample was genotyped using the Illumina Infinium Global Screening-24 (Illumina Inc., USA) at Icahn School of Medicine at Mount Sinai (New York City, USA). The “GenotypeQCtoHRC” module of the DRAGON-data pipeline was used to process the genotype data ^1^. This includes quality control of calls, removing individuals with genotyping rates <0.95, and SNPs with call rates <0.95, minor allele frequency <0.01, and with Hardy-Weinberg Equilibrium mid-*p* < 10^-6^. Imputation and statistical phasing were carried out via the Michigan Imputation Server and minimac4 using the Haplotype Reference Consortium (HRC) panel ^2,3^. Further quality control was performed on the imputed genotypes, described in previous work ^4^. Genetic data, including Polygenic Scores (PGS) for clozapine and norclozapine metabolism, key pharmacogenomic SNPs, and CYP1A2 activity scores, were merged with the combined CLOZUK3 dataset containing FBC and PK information resulting in a dataset of 523 individuals and 1,586 assays.

#### Exploring the effects of CYP1A2 Pharmacogenomic Variation on Neutrophil Counts in Clozapine Users

CYP1A2 plays a key role in clozapine metabolism, therefore it is likely that effects of CYP1A2 pharmacogenomic variability might be observed on neutrophil counts. Pharmacogenomic star alleles for *CYP1A2* were called using PyPGx v0.20.0 ^5^ in Python v3.9.2 ^6^. The imputed genotyping data for CLOZUK3 was passed through the run-chip-pipeline command to derive PGx star alleles for this pharmacogene.

As CPIC-validated information regarding CYP1A2 metabolism phenotypes and activity scores is currently not available, each haplotype was assigned an activity score in line with past work ^7,8^ and described in Supplementary Table 1. These were summed to produce an overall activity score per participant, in which higher scores are reflective of increased enzyme function. Several participants were called with *CYP1A2**1F/1C alleles in the same haplotype. These alleles have opposing functions, therefore in line with past work ^9^, we have treated this haplotype as a normal function allele. Pharmacogenomic variation in *CYP3A4* was not explored despite its key role in clozapine metabolism on account of the rarity of pharmacogenomic star alleles conferring non-normal function ^10^, and likely difficulties that would arise when trying to fit models using such small sample sizes. Likewise, *CYP2D6* variation was not explored as many of the increased function *CYP2D6* pharmacogenomic alleles arise from structural copy number variation that cannot be reliably identified through genotyping arrays ^11^.

The distribution of activity scores is shown in Supplementary Figure 1 and observed allele frequencies compared to previous research in Supplementary Table 1 ^12,13^. The estimated activity scores were included in a linear mixed-effect model alongside medication variables (i.e., daily dose, plasma clozapine and norclozapine concentration), pharmacogenomic SNPs, and other covariates (i.e., age, age^2^, sex, TDS) to determine their impact on ANC.

Finally, two further models were fit to compare the effect size of CYP1A2 activity score with, and without including the intergenic *CYP1A1-CYP1A2* SNP, rs2472297, in the model. There is evidence that this SNP both regulates CYP1A2 activity ^14^ and is in weak linkage disequilibrium with the *1F allele. Therefore, it was added as a covariate in a second model to determine whether its inclusion impacts any association between measures of CYP1A2 activity with ANC in our sample.

#### Assessing the Impact of HLA Genotypes on Neutrophil Counts

Past research has observed associations between HLA genotypes with risk for clozapine-induced agranulocytosis ^15–17^. Therefore, we explored whether these alleles were associated with ANC in the CLOZUK3 sample. Pre-imputation array data was restricted to chromosome 6 using PLINK v1.9 ^18,19^. HIBAG v1.34.1 ^20^ was used to derive HLA alleles, alongside the pre-built InfiniumGlobal-European-HLA4-hg19 prediction model, which best fits the CLOZUK3 data concerning both the genotyping platform and the sample ancestry. This allowed imputation of available HLA genes (-A, -B, -C, -DPB1, -DQA1, -DQB1, -DRB1) to four-digit alleles.

After imputation, alleles with a MAF > 1% were retained for further analysis (Supplementary Table 2), and linear mixed-effect models were used to explore their impact on ANC. Following Levin et al. (2015), genotypes were weighted by posterior imputation probabilities to derive their estimated allele dosage. These were merged with the FBC/PK dataset so that HLA genotypes were associated with the longitudinal assay data (N = 540; 1,627 assays). Each HLA allele was included as a covariate alongside daily clozapine dose, plasma clozapine and norclozapine concentrations, TDS, sex, age, and age^2^. Covariates were standardised and regressed against ANC, with participant ID included as a random intercept term. A total of 116 alleles across 7 HLA genes were included in the regression analyses. Correction for multiple comparisons was performed using the False Discovery Rate (FDR) and Bonferroni correction.

### Supplementary Notes: Statistical Methodology

#### Exclusion of the Metabolic Ratio Variable

Previous work demonstrated that including the clozapine/norclozapine ratio (“metabolic ratio”) as a covariate in a regression model of ANC considerably attenuated the effect sizes of both clozapine and norclozapine plasma concentrations ^22^. However, incorporating ratio variables in regression models has been criticised on the basis that they may result in deceptive statistical artifacts ^23^. Equally, it is possible that the metabolic ratio acts as a collider variable (Supplementary Figure 2; Supplementary Table 3). Controlling for collider variables through their inclusion as covariates, may also introduce spurious associations as described in the causal inference literature ^24^. Therefore, the decision was made to exclude metabolic ratio from the present analyses.

#### R Packages

The full CLOZUK3 dataset was used for linear mixed-effect regression models and single-mediator analyses, benefitting from multiple FBC/pharmacokinetic measurements per patient over time. *lme4* and *lmerTest* were used to fit the LMMs ^25,26^. The mediation package ^27^ was used to perform single-mediation analyses in the longitudinal data.

The CLOZUK3 dataset was transformed into cross-sectional data by taking the lowest observation of ANC for each participant. This reduced dataset was used for multiple-, and single-mediator analyses using Structural Equation Modelling (SEM) in *lavaan* ^28^, and in replication analyses (i.e., Linear Models) of previous research ^22,29^. Before inclusion in all regression and structural equation models, covariates were standardised (mean-centred and scaled) using the *datawizard* R package ^30^.

#### Deriving Residuals for use in Structural Equational Modelling

To account for covariates in the mediation analysis, residualised variables were incorporated during SEM. Each variable was fit as an outcome in separate regression analyses, with the covariates, age, age^2^, sex, and TDS. ANC was included as the outcome in a linear model. Daily clozapine dose was log-transformed and included as an outcome variable in a linear model. Plasma Clozapine and Norclozapine levels were included in a generalised linear model using a gamma distribution and log link function. Linear models were fit using the *lm()* function, and generalised linear models were fit using the *glm()* function. The residuals of the predictors were standardised and then used in place of their parent measure across all mediation analyses unless otherwise specified. In a further model, the impact of CYP1A2 activity score was accounted for by including it alongside age, age^2^, sex, and TDS in the regression models to produce the residualised variables.

### Supplementary Notes: Secondary Analyses

#### Extending the Linear Mixed Models with Genetic Covariates

To assess the impact, if any, of rs2472297 on the association between CYP1A2 activity score with ANC (Supplementary Table 4), two linear mixed-effect models were fit with and without the SNP as an additional covariate in the model. CYP1A2 activity score was significantly associated with ANC (β = 0.145; *p* = 0.004). When the SNP was included as a covariate, there was negligible change in the effect size of CYP1A2 activity score (β = 0.142; *p* = 0.011), and as in the previous pharmacogenomic analysis (Table 3, Main Text), rs2472297 was not associated with ANC in this model (β = 0.004; *p* = 0.949). This indicates that the observed association between CYP1A2 activity score and ANC is likely not conflated with rs2472297.

There was no evidence of associations between Polygenic Scores for either clozapine metabolism (β = 0.034; *p* = 0.532) or norclozapine metabolism (β = -0.009; *p* = 0.863) with ANC. However, a strong negative association (β = -0.770; *p* = 0.002) between the Duffy-Null genotype and ANC was observed in the CLOZUK3 sample. The presence of the Duffy-Null genotype was linked with a decline in neutrophils of approximately 770 cells/mm^3^ in comparison to non-carriers. Full estimates from these additional genetic models are included in Supplementary Table 5.

HLA genotypes were similarly included in linear mixed-effect models to explore whether variation in the HLA region influenced neutrophil levels in clozapine users. An overview of these findings is shown in Supplementary Table 6. After controlling for multiple comparisons, no HLA alleles were significantly associated with ANC in the CLOZUK3 sample; this suggests little influence of the HLA region on neutrophil counts in these participants. Prior to this correction, 3 HLA alleles demonstrated nominally significant associations. These included *HLA-DRB1*16:01* (β = 0.087; *p* = 0.001) and *HLA-DRB1*04:04* (β = 0.073; *p* = 0.011), which were positively associated with ANC, alongside *HLA-DRB1*01:03* (β = -0.068; *p* = 0.016), which was inversely associated with ANC.

#### Controlling for CYP1A2 Activity in the Mediation Analysis

Linear mixed-effect models revealed a significant association between CYP1A2 activity score and ANC in the CLOZUK3 sample. Thus, to account for the impact of this pharmacogenomic variation on clozapine metabolism, and potentially neutrophil levels, the activity score was residualised out of the included variables, as previously described. In this instance, both the association between dose and ANC (β = 0.100, *p* = 0.077), and the indirect effect via clozapine and norclozapine were weakened (β = 0.050, *p* = 0.077). While this attenuation of effects may be due to the inclusion of the CYP1A2 activity score, it could also arise because of the reduced sample size with complete genetic information. Therefore, the primary model was repeated on this subset of the CLOZUK3 sample, to determine how this smaller sample size affected the model output. This resulted in inflated estimates for the previously significant direct (β = 0.251, *p* = 0.077) and indirect effects (β = 0.125, *p* = 0.077). A comparison of the three models (Supplementary Figure 3) showed that using a reduced sample size results in larger confidence intervals indicative of a loss of statistical power. Interestingly, controlling for CYP1A2 activity score reduces this uncertainty, bringing the intervals in line with those seen in the primary model, albeit with slightly smaller effect sizes.

#### Non-Residualised SEM for Mediation Analysis

SEM was also used on non-residualised versions of the variables (Supplementary Figure 4) to ensure that controlling for covariates in this way didn’t create spurious results. As in the residualised model, there was a significant positive effect of daily clozapine dose on lowest ANC (β = 0.110, *p* = 0.017). Equally, there was a significant indirect effect via both clozapine and norclozapine plasma concentrations (β = 0.054, *p* = 0.015) with no evidence for mediation by plasma clozapine concentration alone (β = - 0.017, *p* = 0.254).

#### Single Variable Mediation Analyses

SEM was performed with residualised plasma clozapine concentration as the lone mediator between dose and lowest ANC (Supplementary Table 7). As before, a significant effect of dose on lowest ANC was observed (β = 0.172, *p* = 1.1 x 10^-4^), with no support for clozapine as a mediating variable (β = 0.008, *p* = 0.394). A secondary, causal mediation analysis was performed using the full CLOZUK3 dataset. These results are consistent with the findings derived from using SEM, showing a significant direct effect of daily clozapine dose (ADE = 0.155, *p* < 2 × 10^-16^) and no indirect effect transmitted via clozapine plasma concentration (ACME = 0.006, *p* = 0.574).

A complementary single-mediator SEM was performed with norclozapine as the sole mediating variable. A significant direct effect of daily clozapine dose on lowest ANC was observed (β = 0.154, *p* = 6.1 x 10^-4^). Additionally, there is some evidence of a mediating effect when we consider norclozapine plasma concentration alone (β = 0.025, *p* = 0.048). When this analysis was replicated using the longitudinal sample, a significant direct effect was observed again (ADE = 0.133, *p* < 2 × 10^-16^). Furthermore, a significant indirect effect via norclozapine was also observed (ACME = 0.029, *p* = 0.008), shown in Supplementary Table 8.

#### Replicating Past Studies with Linear Models.

Three linear models were fit in-line with Willcocks et al., (2021), excluding the covariates Time on Treatment (absent in CLOZUK3) and Days between Assays (as FBC and pharmacokinetic assays were performed on the same day for all our data points), with results reported in Supplementary Table 9. A forest plot comparing standardised regression coefficients obtained from the CLOZUK3 and CLOZUK2 samples is shown in Supplementary Figure 5.

In the first model, daily dose (β = 0.126; *p* = 0.006) and norclozapine level (β = 0.208; *p* = 0.015) were both positively associated with ANC. Although not significant, clozapine was negatively associated with ANC (β = -0.121; *p* = 0.149). The addition of metabolic ratio in the second model, reduced effect sizes for both clozapine plasma concentration (β = -0.052; *p* =0.729) and norclozapine plasma concentration (β = 0.139; *p* = 0.358). Daily clozapine dose was unaffected by this (β = 0.127; *p* = 0.006), and the metabolic ratio itself was not significantly associated with ANC (β = -0.045; *p* = 0.581). In the final model, no pharmacogenomic SNPs were associated with ANC. Furthermore, the previous association between daily dose and ANC became smaller and non-significant (β = 0.073; *p* = 0.215).

### Supplementary Figures

#### Supplementary Figure 1 – Distribution of CYP1A2 Activity Scores in CLOZUK3

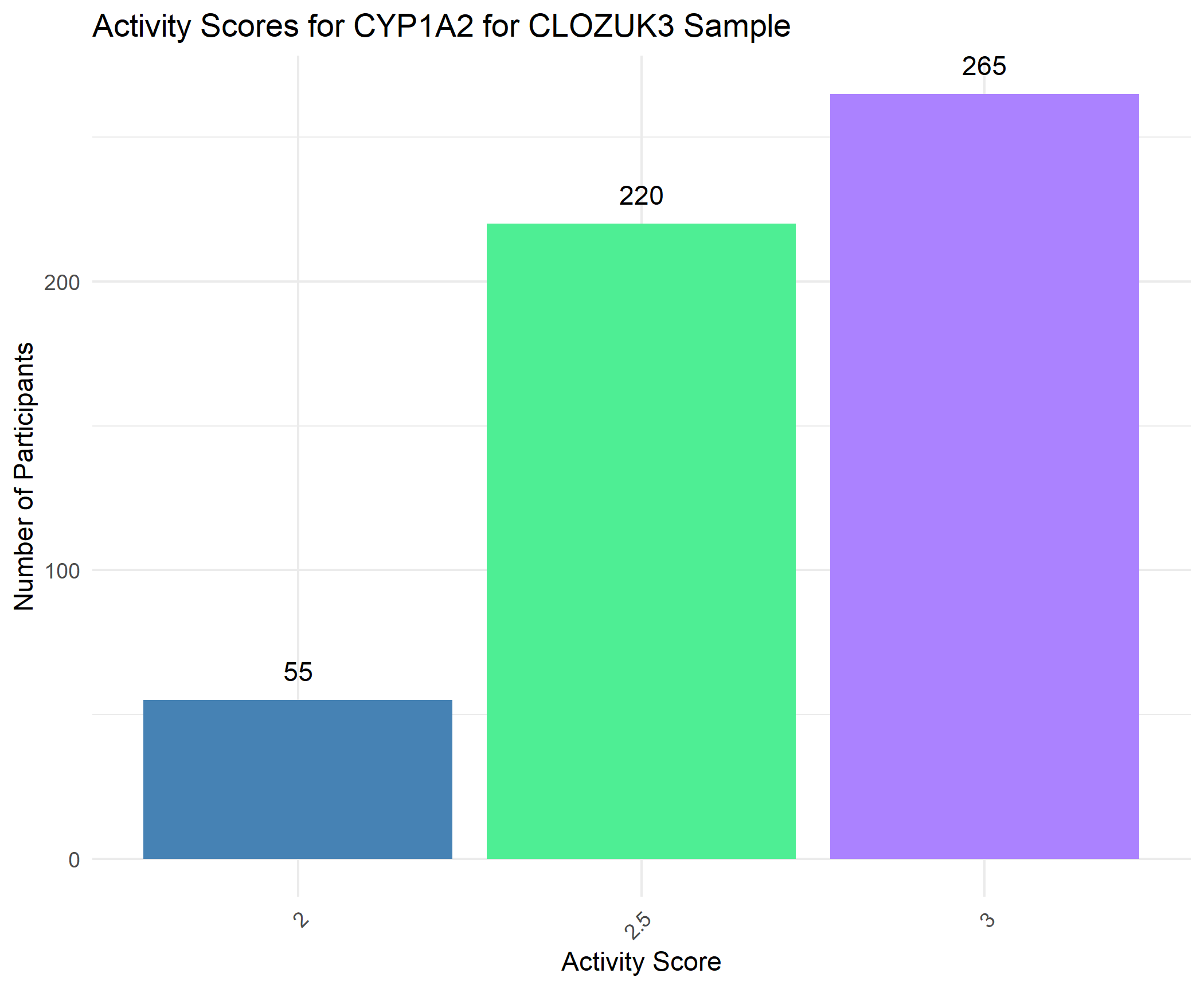

Supplementary Figure 1. Distribution of pharmacogenomic allele derived CYP1A2 activity scores observed in the CLOZUK3 sample.

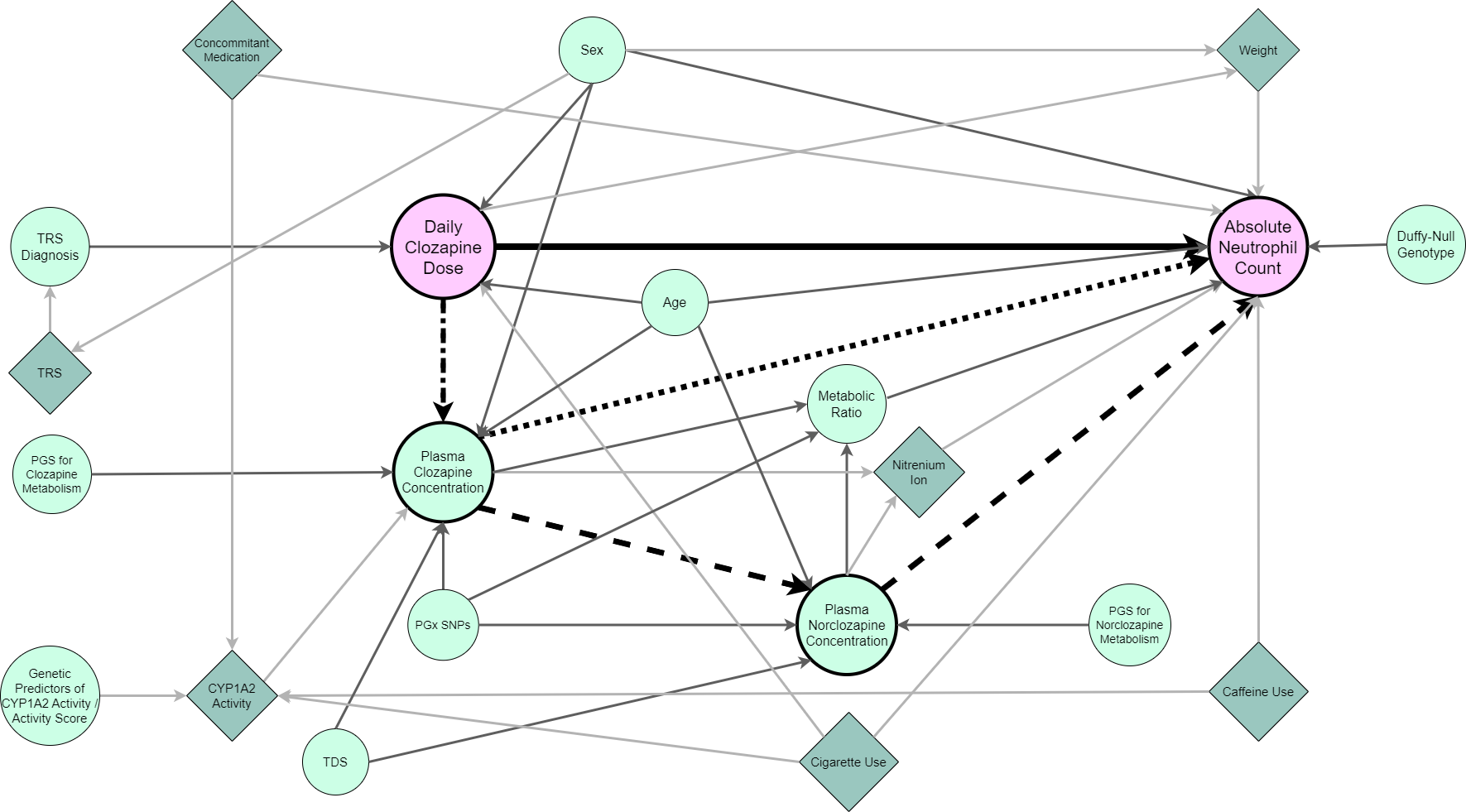

#### Supplementary Figure 2 – Directed Acyclic Graph showing causal paths between Clozapine Dose and Absolute Neutrophil Count.

Supplementary Figure 2. Variables included in Directed Acyclic Graph showing the possible causal associations between clozapine dose and absolute neutrophil count. Pink circles represent the exposure (Daily Clozapine Dose) and outcome (Absolute Neutrophil). Light green circles represent measured variables, whereas dark green diamonds represent latent variables. Dark grey arrows represent paths between observed variables, whereas light grey arrows represent any path in which one (or both) of the variables involved are unobserved. Black paths represent effects of interest in the main regression and mediation analyses (solid = direct effect; dotted = indirect effect via mediator 1; dashed = indirect effect via mediators 1 & 2; dot-dash = path shared by both indirect effects). TRS = Treatment Resistant Schizophrenia; PGS = Polygenic Score; PGx = Pharmacogenomic; TDS= Time between Dose and Sample.

#### Supplementary Figure 3 – Comparison of Mediation Analysis Effect Sizes with and without including Genetic data.

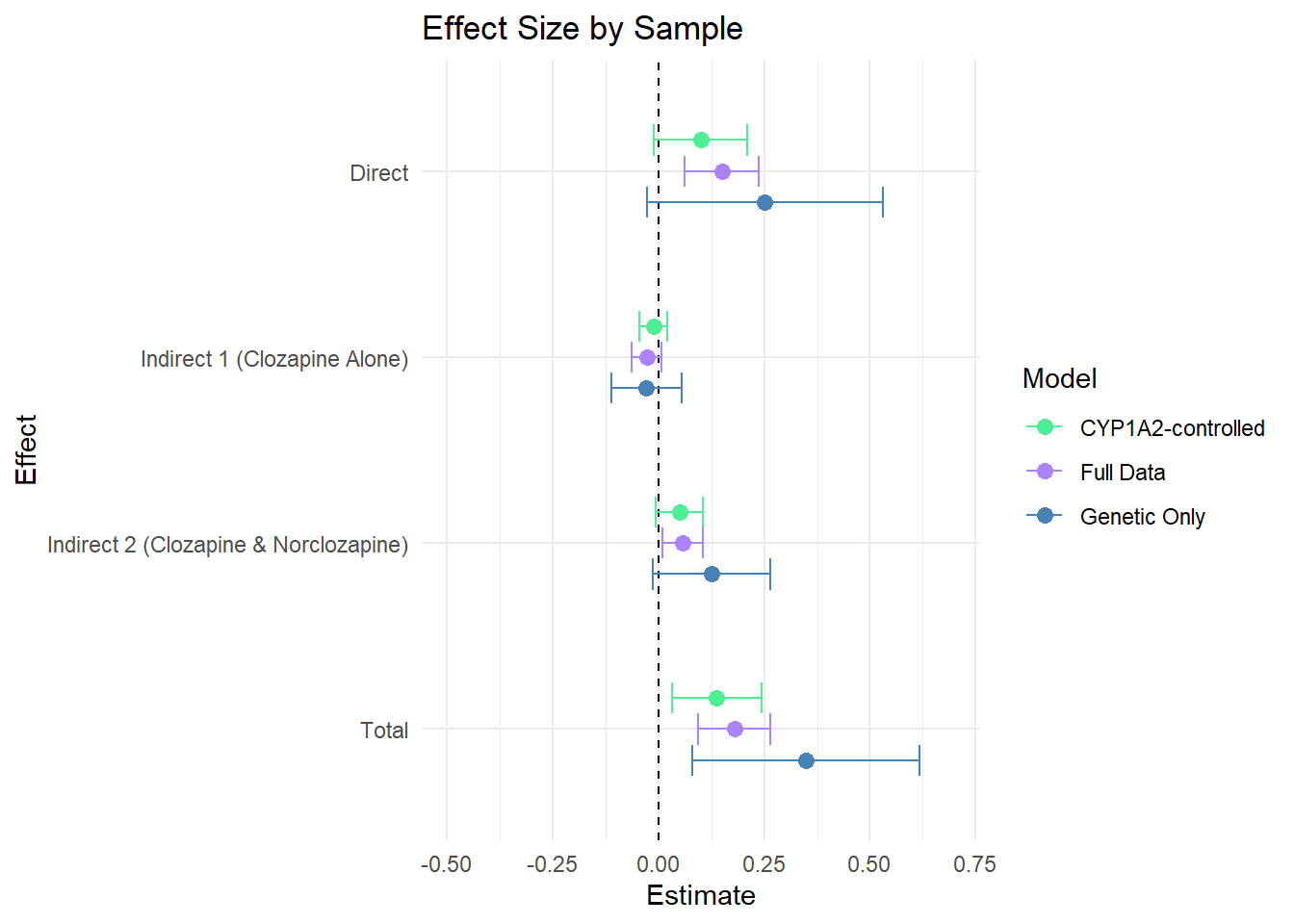

Supplementary Figure 3. Forest plot comparing effect estimates between the three Structural Equation Models. ‘Full data’ refers to the first model containing the full CLOZUK3 sample. ‘Genetic Only’ refers to the same model as in Full data, but performed on the subset of the sample for whom genetic data is available. The final model, ‘CYP1A2-controlled’, was performed on the genetic only sample but controls for CYP1A2 activity scores when creating the residualised variables. Effect estimates are standardised, and error bars show 95% confidence intervals.

#### Supplementary Figure 4 – Results of Mediation Analysis using SEM with non-residualised variables.

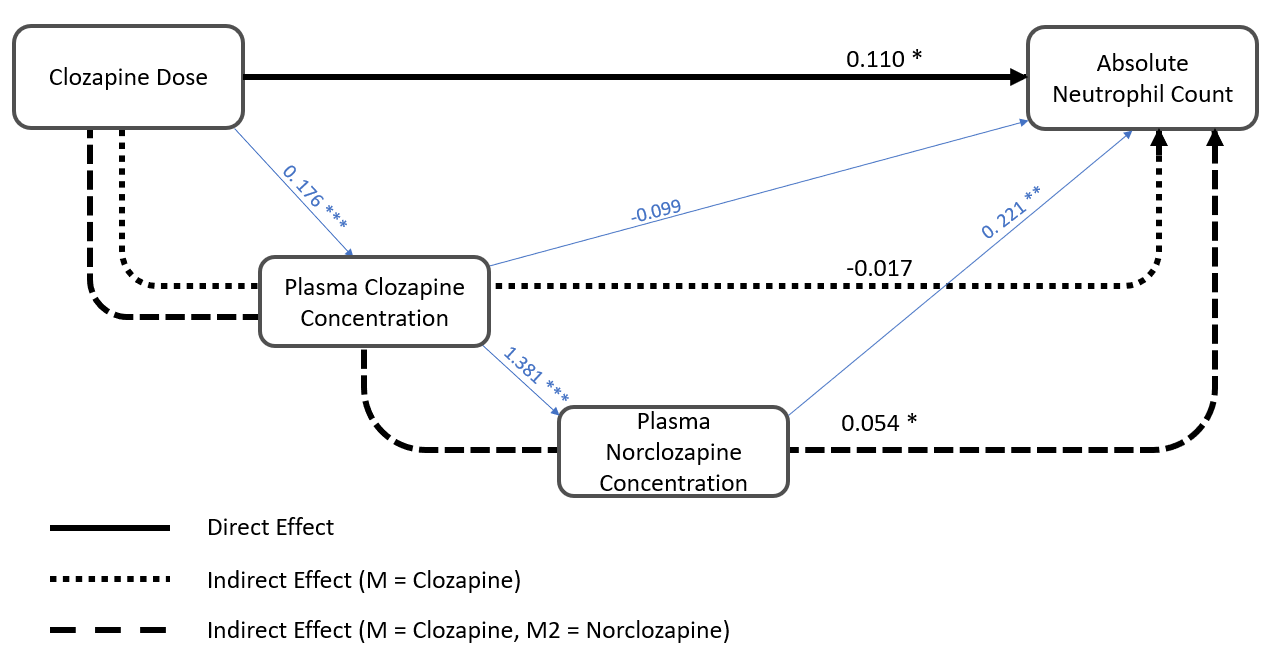

Supplementary Figure 4. Path diagram showing association between Dose and Lowest ANC with Clozapine and Norclozapine as mediators. Plot edges are labelled with standardised regression coefficients. Variables included in SEM are the parent, non-residualised variables. SEM = Structural Equation Modelling.

* p<0.05	** p<0.01	*** p<0.001

#### Supplementary Figure 5 – Comparison of Covariate Effect Sizes from Linear and Linear Mixed Effect Models on CLOZUK3 with Linear Model on CLOZUK2.

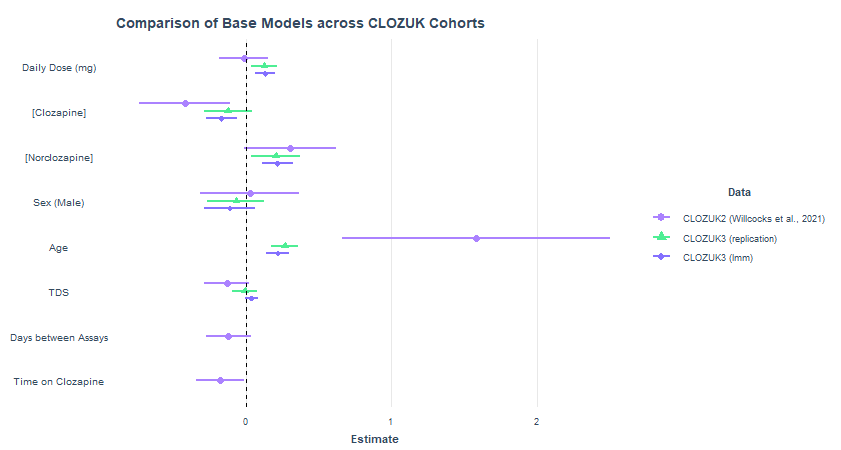

Supplementary Figure 5. Forest plot comparing standardised regression coefficients from equivalent models between CLOZUK2 (Willcocks et al., 2021) and CLOZUK3 (present work) datasets. The CLOZUK2 and CLOZUK3 (replication) models explore the association between covariates and lowest value of ANC in cross-sectional samples. The CLOZUK3 (lmm) model utilises the available longitudinal data, incorporating a random effect variable for participant ID. Error bars show 95% Confidence Intervals. ANC = Absolute Neutrophil Count; TDS = Time Between Dose and Sample; lmm = Linear Mixed-Effect Model.
